# A Multi-State Markov Model for the Longitudinal Analysis of Clinical Composite Outcomes in Heart Failure

**DOI:** 10.1101/2023.12.05.23299570

**Authors:** David Lora Pablos, Andrea Leiva-García, José Luis Bernal, Jorge Vélez, Beatriz Palacios, Miriam Villarreal, Margarita Capel, Nicolás Rosillo, Miguel Hernández, Héctor Bueno

## Abstract

**Background:** The statistical analysis of composite outcomes is challenging. The Clinical Outcomes, HEalthcare REsource utilizatioN, and relaTed costs (COHERENT) model was developed to describe and compare all components (incidence, timing and duration) of composite outcomes, but its statistical analysis remained unsolved. The aim of the study is to assess a multi-State Markov model as one statistical solution for the COHERENT model.

**Methods:** A cohort of 3280 patients admitted to the emergency department or hospital for heart failure during year 2018 were followed during one year. The state of the patient was registered at the end of each day during 365 days as: home, emergency department (ED), hospital, re-hospital, re-ED, and death. Outcomes of patients with or without severe renal disease (sRD) were compared as an example. A Multi-State Markov model was developed to explain transitions to and from these states during follow-up.

**Results:** A Multi-State Markov model showed, adjusted for age and sex, a significantly lower likelihood of patients with sRD to return home regardless of the state in which they were (ED → HOME (HR, 0.72; 95%CI, 0.54-0.95), RE-ED → HOME (HR, 0.83; 95%CI, 0.75-0.93), HOSPITAL → HOME (HR, 0.77; 95%CI, 0.69-0.86), RE-HOSPITAL → HOME (HR, 0.82; 95%CI, 0.74-0.92) and a higher mortality risk, in particular at the hospital and at home (HOME → Death [HR, 1.54; 95%CI, 1.01-2.37] and HOSPITAL → Death [HR, 1.71; 95%CI, 1.30-2.24].

**Conclusion:** Multi-state Markov models offer a statistical solution for the comprehensive analysis of composite outcomes assessed as transitions from different clinical states.

**Clinical Perspective:**

- What is new?
  - An integrated analysis of all components of composite endpoints including its incidence and duration is possible using the COHERENT model with analysis of transition risks.
  - A statistical approach based on Markov chain models is a new potential statistical solution for the multivariate estimation of the risk of transitions in mutually exclusive composite endpoints.
- What are the clinical implications?
  - The use of the COHERENT model and Markov models is an opportunity to analyze composite endpoints and understand better the relationships between its components and, potentially, to improve the performance of statistical analysis in randomized controlled trials.
  - The utilization of the COHERENT model and Markov models in randomized controlled trials should be validated in future observational studies and in randomized controlled trials.

## Introduction

Heart failure (HF) is a good scenario for the use of composite outcomes^1–5^, where mortality and readmissions have been frequently used as a composite for evaluating efficacy in clinical randomized trials. The relevance of outcome analysis in HF is obvious given that it is a major health problem with most patients with HF having at least one hospitalization during their lives, and several having subsequent readmissions, causing a huge burden on health systems and economies. We recently developed the Clinical Outcomes, HEalthcare REsource utilizatioN, and relaTed costs (COHERENT) model, a new approach for presenting and analysing mutually exclusive endpoints, including clinical endpoints, resource utilization and costs over time in a visual way^6,7^. However, the statistical approach for such complex amount of information has not been solved so far^6,7^

Clinical outcomes may be understood as states within the evolution of the disease. For instance, hospitalization, discharge or readmission, being death the final outcome. These states may or may not be visited by the patients during their clinical journey. Some authors have proposed to explain the clinical course of chronic diseases as a multi state model where outcomes can be understood as transient or intermediate events with a final outcome. HF may be explained as a multistate model^8^ which is a Markov process under the assumption that the chance of entering a new state at the start of each cycle does not depend on the path taken by the individual to reach the current state but on exposure factors and the risk of transition from one state to another. A number of authors have extended the concepts of multi-state Markov models to longitudinal data, where the time spent in each state was incorporated, generating the multi-state semi-Markov model^9,10^, or incorporating time-dependent explanatory variables^11^

We present here a simple statistical solution using the multi-state Markov model for longitudinal data for the statistical analysis of multiple clinical endpoints over time to be used with the COHERENT model^6^.

## Methods

### Population

The description of the population used for this study has been presented elsewhere^7^. In brief, 3280 patients with HF who were seen in the emergency department (ED) and/or were hospitalized in a tertiary hospital with a primary or secondary diagnosis of HF during one calendar year (2018) were included. Patients were then followed up for one year.

### Multi-state model of longitudinal data

The process of clinical care for patients with HF was conceptualized using the first visit to the ED as the initial state, and from then the other clinical outcomes were defined and classified as intermediate events (home, re-ED, hospitalization, re-hospitalization) and the final state (death). This is described though a multi-state model of longitudinal data based on the Markov model on longitudinal data^8,12,13^. The Markov process refers to the assumption that the chance of entering a new state at the start of each cycle does not depend on the path the individual took to their current state (although the chance may depend on the cycle and other risk factors). The death state is considered as an absorbing state, that is, the individual cannot move out from this state.

The state of the patient was registered at the end of each day during the follow-up period (one year in this case) with the exception of ED stays, which accounted for a minimum of one full day if the patient was discharged home on the same day regardless of the time staying in the ED on that particular day, meaning that information of the state between days was not available. Therefore, one patient could stay in more than one state in the same day. Transitions were defined as illustrated in **Figure 1**. Direct transitions between ED and re-ED and between hospital and re-hospital were not counted.

**Figure 1.**
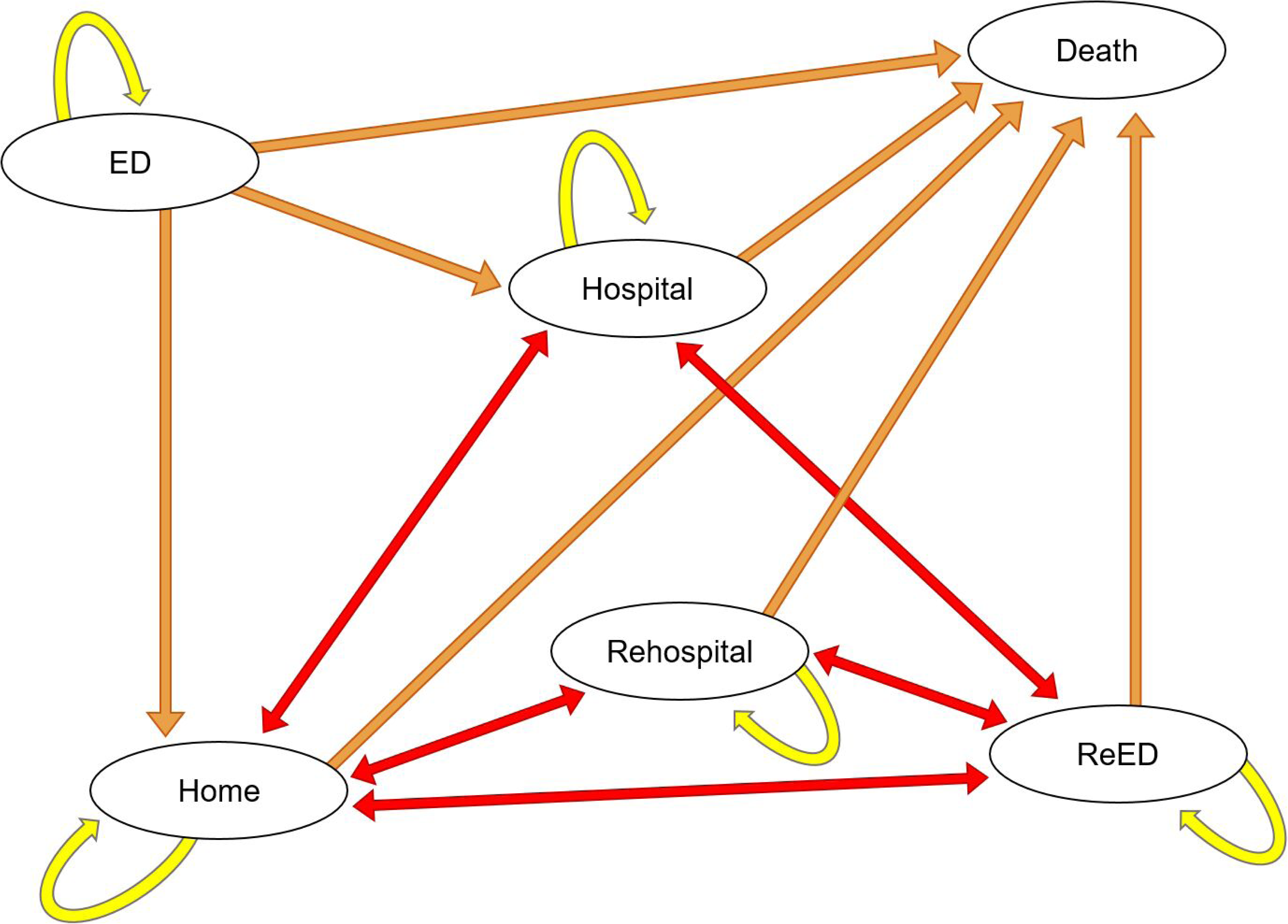
States in the Markov chain used in the analysis for the current COHERENT model. **Abbreviations.** ED (initial state), Hospital, Rehospital, Home, ReED, and Death (final state). Yellow arrows indicate the permanence in the same state, Orange arrows represent forward transitions and red arrows forward and backward transitions

We used Xt to indicate the state space occupied by the individual at time t, with t=0 … 365 days and state space = {ED, Hospital, Home, Re-hospital, Re-ED, and Death}. The matrix of transition probabilities for the Markov process, (Xt)t≥0, was defined as:

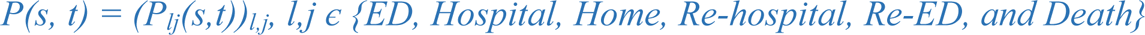

with transition probabilities:

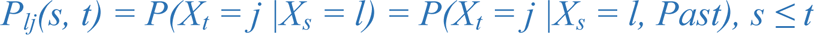

and definition of the transition hazards in the Markov process with l -> j transition at t as:

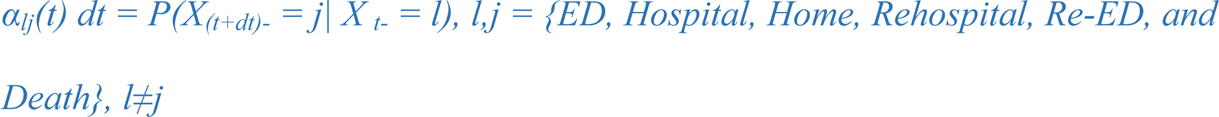

The Markov proportional hazards model ^2,14,15^ was used to evaluate a common effect of one covariate as an example. In this particular case we used severe renal disease (sRD)—defined as the presence of baseline glomerular filtration rate values <30 ml/min/1.73 m^2^ or serum creatinine values >2 mg/dL if baseline glomerular filtration rate was not available, a well known predictor or poorer outcomes in patients with HF ^16,17^— as the variable of interest to test the model with the following model:

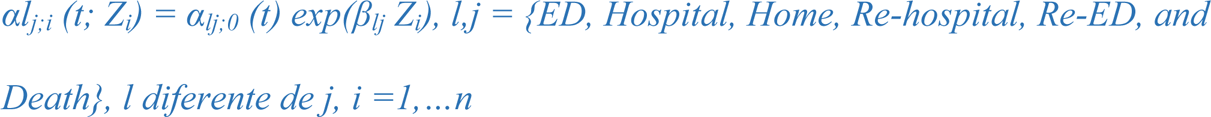

where β_lj_ is a 1 × p vector of regression coefficients, Z_i_ is a p × 1 vector of covariates for individual i, and α_lj;0_(t) is an unspecified, non-negative baseline hazard function. Hazard ratio (HR) estimates are presented with 95% confidence intervals (95%CI). All calculations were done using a R^18^ package msm^19^.

## Results

A total of 3280 patients with HF were included, mean age 80.9 years (11.3), 1830 (55.8%) women. Of these, 496 patients (15.1%) had severe renal disease. Baseline characteristics according to the presence of severe renal disease are presented in **Table 1**. Patients with sRD were older, more frequently women and had more comorbidities. Patient with sRD presented also worse outcomes at 30 days and 1 year (**Table 2** and **Figure 2**).

**Figure 2.**
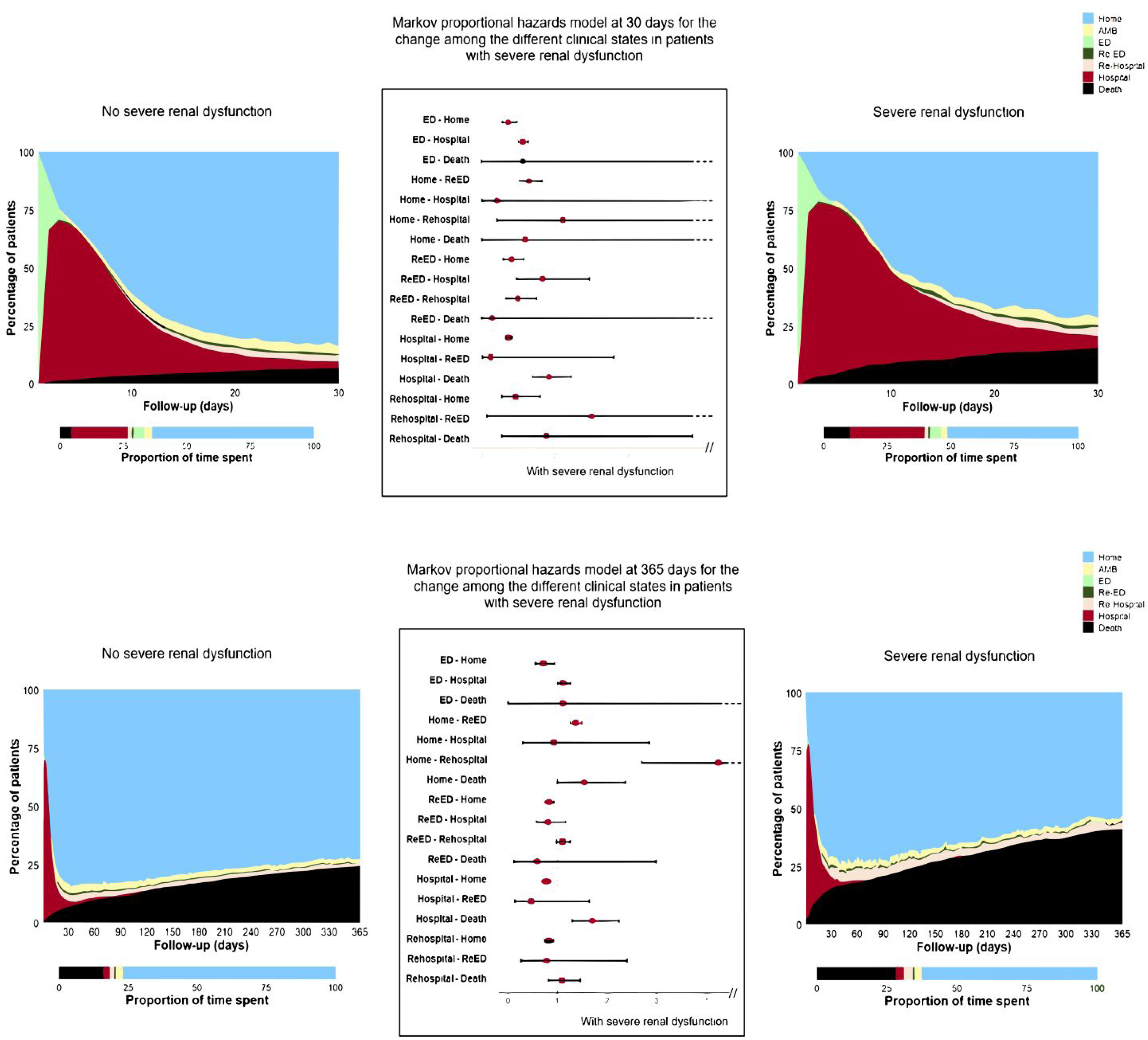
Composite outcomes graph using the COHERENT model comparing patients with severe renal dysfunction (right) and patients without severe renal dysfunction (left) using the Markov proportional hazard models. **A. 30 days follow-up** **2.B. 1 year follow-up**

**Table 1.** Baseline characteristics according to the presence of severe renal dysfunction.

|  | No severe renal disease<br>(n=2784) | Severe renal disease<br>(n=496) | P value |
| --- | --- | --- | --- |
| **Age** | 83.0 (75.0 - 88.0) | 85.0 (78.0 - 89.0) | <0.001 |
| **Age group** |  |  | 0.002 |
| Younger than 65 | 285 (10.24%) | 35 (7.06%) |  |
| Between 65 and 74 | 357 (12.82%) | 56 (11.29%) |  |
| Between 75 and 84 | 898 (32.26%) | 139 (28.02%) |  |
| Older than 85 | 1244 (44.68%) | 266 (53.63%) |  |
| **Female sex** | 1557 (55.93%) | 273 (55.04%) | 0.751 |
| **Diabetes** | 1153 (41.41%) | 275 (55.44%) | <0.001 |
| **AHT** |  |  | 0.410 |
| Hipertension | 1020 (36.63%) | 200 (40.32%) |  |
| Hipotension | 39 (1.40%) | 7 (1.41%) |  |
| Normotension | 1651 (59.30%) | 279 (56.25%) |  |
| Not available data | 74 (2.66%) | 10 (2.02%) |  |
| **Heart rate** |  |  | 0.012 |
| High heart rate | 378 (13.58%) | 48 (9.68%) |  |
| Low heart rate | 257 (9.23%) | 63 (12.70%) |  |
| Normal heart rate | 2075 (74.53%) | 375 (75.60%) |  |
| Not available data | 74 (2.66%) | 10 (2.02%) |  |
| **Smoking** | 941 (33.80%) | 154 (31.05%) | 0.252 |
| **Dyslipidaemia** | 1271 (45.66%) | 147 (29.64%) | <0.001 |
| **LVEF** |  |  | 0.050 |
| HFmrEF | 230 (8.26%) | 37 (7.46%) |  |
| HFpEF | 1869 (67.13%) | 349 (70.36%) |  |
| HFrEF | 293 (10.52%) | 59 (11.89%) |  |
| HF with unknown LVEF | 69 (2.48%) | 15 (3.02%) |  |
| Not available echocardiogram | 323 (11.60%) | 36 (7.26%) |  |
| **COPD** | 242 (8.69%) | 32 (6.45%) | 0.116 |
| **Cancer** | 115 (4.13%) | 19 (3.83%) | 0.851 |
| **Hypertensive Heart Disease** | 373 (13.40%) | 47 (9.48%) | 0.020 |
| **Cerebrovascular disease** | 7 (0.25%) | 1 (0.20%) | 1.000 |
| **Atrial fibrillation** | 641 (23.02%) | 96 (19.35%) | 0.081 |
| **Heart valve disease** | 197 (7.08%) | 24 (4.84%) | 0.083 |
| **Dementia** | 11 (0.40%) | 2 (0.40%) | 1.000 |
| **Respiratory insufficiency** | 1023 (36.74%) | 169 (34.07%) | 0.276 |
| **Ischemic Heart Disease** | 414 (14.87%) | 76 (15.32%) | 0.848 |

**Table 2.** Clinical outcomes according to the presence of several renal dysfunction.

|  | **No severe**<br>**renal disease**<br>(n=2784) | **Severe**<br>**renal disease**<br>(n=496) | **P value** |
| --- | --- | --- | --- |
| **30-day mortality** | 191 (6.86%) | 77 (15.52%) | <0.001 |
| **365-day mortality** | 677 (24.32%) | 204 (41.13%) | <0.001 |
| **In-hospital mortality** | 145 (5.21%) | 63 (12.70%) | <0.001 |
| **30-day admissions** | 269 (9.66%) | 58 (11.69%) | 0.190 |
| **365-day admissions** | 886 (31.82%) | 209 (42.13%) | <0.001 |
| **30-day readmissions** | 77 (2.77%) | 20 (4.03%) | 0.165 |
| **365-day readmissions** | 360 (12.93%) | 60 (12.09%) | 0.660 |
| **30-day re-ED visits** | 501 (17.99%) | 99 (19.96%) | 0.327 |
| **365-day re-ED visits** | 1717 (61.67%) | 314 (63.30%) | 0.522 |

Of these, 880 (26.8%) patients died during follow-up, 204 (41.13%) with sRD y 677 (24.28%) without sRD (RR, 1.69; 95%CI, 1.50-1.92; p<0.001). Among all patients, only 247 (7.53%) had only one transition (41 died in the ED and 206 were discharged home directly and had no further state transitions) while 818 patients (24.9%) had >7 changes of state, of whom 197 had died at the end of follow-up. No statistical differences were found between the number of state transitions and death.

Patients with sRD spent a lower number of days at home 98.3 (50.5; 177.0) compared with those without sRD (147.0 [76.5; 348.0]; p <0.001). They also spent more time in re-ED and in hospital (8.0 [5.0, 15.0] vs. 7.0 [5.0, 11.0] days) p<0.001. The times spent in each clinical state by the presence of sRD are shown in **Supplementary Table 1**)

**Table 3** presents the results of the analysis for sRD with the crude model (left column) and an age- and sex-adjusted model (three right columns). Both models confirm the significantly lower likelihood of patients with sRD to return home regardless of the state in which they were (ED → HOME (HR, 0.72; 95%CI, 0.54-0.95), RE-ED → HOME (HR, 0.83; 95%CI, 0.75-0.93), HOSPITAL → HOME (HR, 0.77; 95%CI, 0.69-0.86), RE-HOSPITAL → HOME (HR, 0.82; 95%CI, 0.74-0.92) and their higher mortality risk, in particular at the hospital and at home (HOME → Death [HR, 1.54; 95%CI, 1.01-2.37] and HOSPITAL → Death [HR, 1.71; 95%CI, 1.30-2.24] adjusted for age and sex. Additionally, patients with sRD have a higher likelihood of return to the ED (HR, 1.37; 95%CI, 1.26-1.49) or being re-hospitalized (HR, 4.25; 95%CI, 2.70-6.70).

**Table 3.**
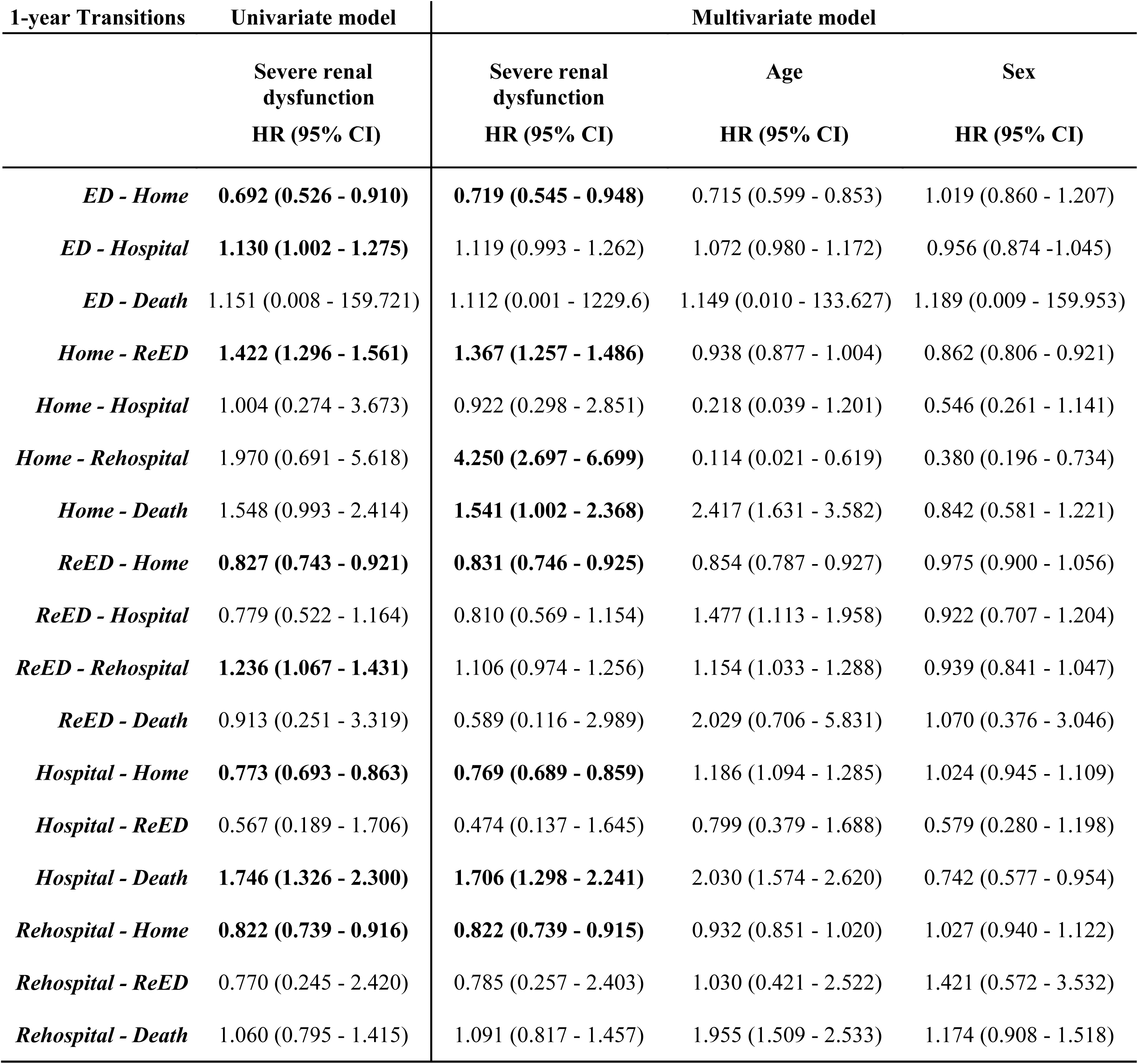
Markov proportional hazards model at 1 year for the change among the different clinical states in patients with severe renal dysfunction (univariate model) and Markov proportional hazards model with severe renal dysfunction as the variable of interest adjusted for age and sex (multivariate model)

| 1-year Transitions | Univariate model | Multivariate model |  |  |
| --- | --- | --- | --- | --- |
|  | Severe renal dysfunction<br>HR (95% CI) | Severe renal dysfunction<br>HR (95% CI) | Age<br>HR (95% CI) | Sex<br>HR (95% CI) |
| *ED - Home* | **0.692 (0.526 - 0.910)** | **0.719 (0.545 - 0.948)** | 0.715 (0.599 - 0.853) | 1.019 (0.860 - 1.207) |
| *ED - Hospital* | **1.130 (1.002 - 1.275)** | 1.119 (0.993 - 1.262) | 1.072 (0.980 - 1.172) | 0.956 (0.874 - 1.045) |
| *ED - Death* | 1.151 (0.008 - 159.721) | 1.112 (0.001 - 1229.6) | 1.149 (0.010 - 133.627) | 1.189 (0.009 - 159.953) |
| *Home - ReED* | **1.422 (1.296 - 1.561)** | **1.367 (1.257 - 1.486)** | 0.938 (0.877 - 1.004) | 0.862 (0.806 - 0.921) |
| *Home - Hospital* | 1.004 (0.274 - 3.673) | 0.922 (0.298 - 2.851) | 0.218 (0.039 - 1.201) | 0.546 (0.261 - 1.141) |
| *Home - Rehospital* | 1.970 (0.691 - 5.618) | **4.250 (2.697 - 6.699)** | 0.114 (0.021 - 0.619) | 0.380 (0.196 - 0.734) |
| *Home - Death* | 1.548 (0.993 - 2.414) | **1.541 (1.002 - 2.368)** | 2.417 (1.631 - 3.582) | 0.842 (0.581 - 1.221) |
| *ReED - Home* | **0.827 (0.743 - 0.921)** | **0.831 (0.746 - 0.925)** | 0.854 (0.787 - 0.927) | 0.975 (0.900 - 1.056) |
| *ReED - Hospital* | 0.779 (0.522 - 1.164) | 0.810 (0.569 - 1.154) | 1.477 (1.113 - 1.958) | 0.922 (0.707 - 1.204) |
| *ReED - Rehospital* | **1.236 (1.067 - 1.431)** | 1.106 (0.974 - 1.256) | 1.154 (1.033 - 1.288) | 0.939 (0.841 - 1.047) |
| *ReED - Death* | 0.913 (0.251 - 3.319) | 0.589 (0.116 - 2.989) | 2.029 (0.706 - 5.831) | 1.070 (0.376 - 3.046) |
| *Hospital - Home* | **0.773 (0.693 - 0.863)** | **0.769 (0.689 - 0.859)** | 1.186 (1.094 - 1.285) | 1.024 (0.945 - 1.109) |
| *Hospital - ReED* | 0.567 (0.189 - 1.706) | 0.474 (0.137 - 1.645) | 0.799 (0.379 - 1.688) | 0.579 (0.280 - 1.198) |
| *Hospital - Death* | **1.746 (1.326 - 2.300)** | **1.706 (1.298 - 2.241)** | 2.030 (1.574 - 2.620) | 0.742 (0.577 - 0.954) |
| *Rehospital - Home* | **0.822 (0.739 - 0.916)** | **0.822 (0.739 - 0.915)** | 0.932 (0.851 - 1.020) | 1.027 (0.940 - 1.122) |
| *Rehospital - ReED* | 0.770 (0.245 - 2.420) | 0.785 (0.257 - 2.403) | 1.030 (0.421 - 2.522) | 1.421 (0.572 - 3.532) |
| *Rehospital - Death* | 1.060 (0.795 - 1.415) | 1.091 (0.817 - 1.457) | 1.955 (1.509 - 2.533) | 1.174 (0.908 - 1.518) |

The multi-state models were tested for 30-day (**Supplementary Table 2**) and 1-year outcomes (**Table 3**). **Figure 3** shows the effect of severe renal dysfunction on the transition risks from one to other states at 30 days and 365 days.

**Figure 3.**
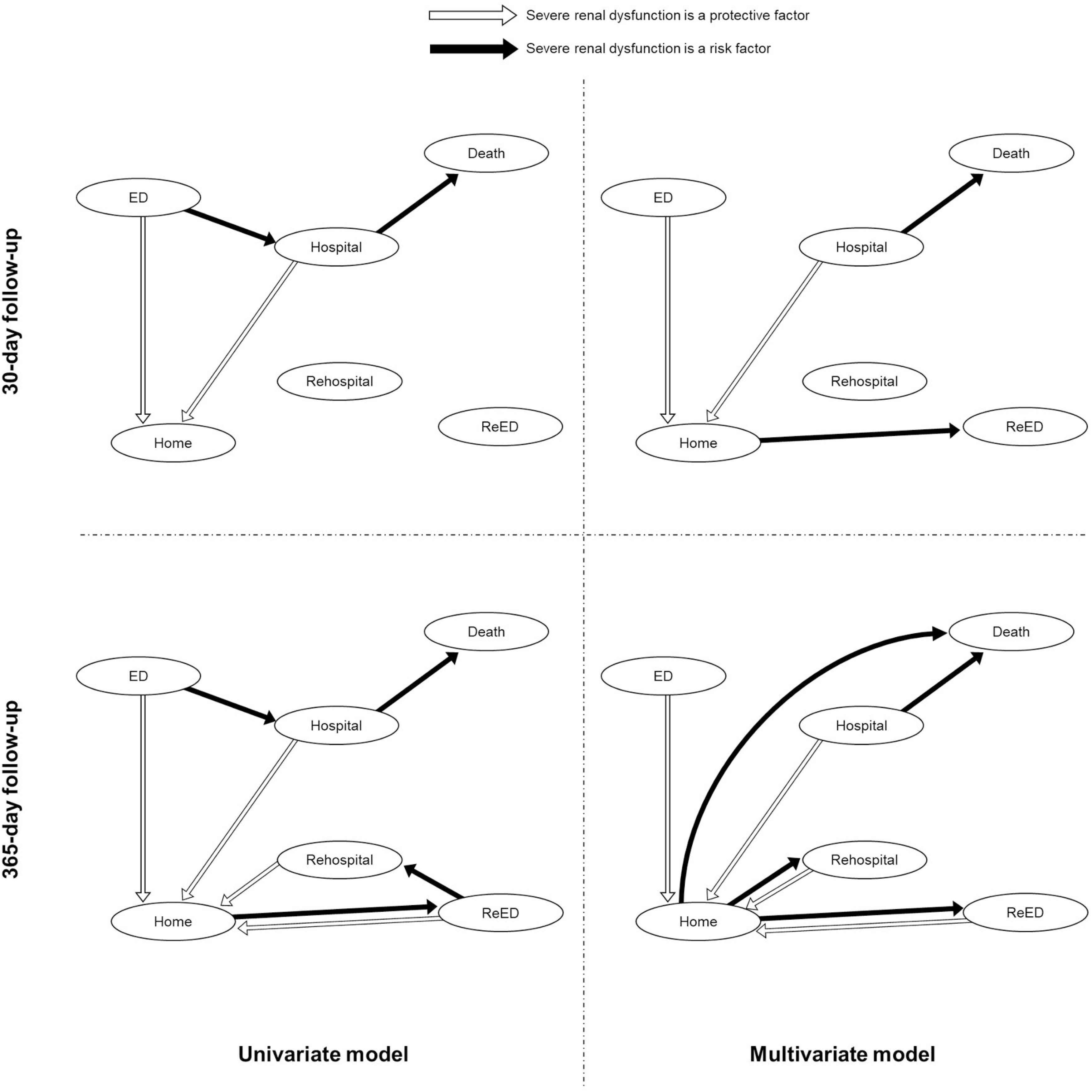
Significant changes of state in the Markov chain used in the analysis for 30 and 365-day follow up.

## Discussion

We have applied a statistical approach for the analysis of multiple composite outcomes over time based on multi-state Markov models for longitudinal data that can be used with the COHERENT model. This approach allows the calculation of the probabilities of transitions available between all the components of the mutually exclusive endpoints contained in the model and the study of the effect of covariables on the probability (risk) of transitioning from one to other state. Our model is, therefore, an attempt to improve the complex analysis of composite endpoints over time when these include multiple components with different incidences and durations, and are registered with different times of initiation and termination, and potential censoring in follow-ups. As we acknowledged in the presentation of the COHERENT model the statistical approach for these analyses is quite complex and had not been solved so far^6^.

Multistate Markov models improve current competing risk models providing not only with the risks of the transitions between the components of interest of the composite endpoints (i.e. home, hospital, re-hospitalization, re-ED visit, and the absorbing state, death), facilitating the description of the probabilities of the patientś trajectories^8,11^. Thus, multistate Markov Models allow describing the risks and the related costs of events occurring after the index contact (ED visit in this particular case).

This method allows multivariate analysis, estimating the risk of transitioning from one state to any other one as a function of other covariates, such as baseline characteristics, providing with adjusted hazard ratios for each transition as shown in the results. In the example, the presence of sRD is associated with an increased risk of in-hospital and out of hospital death with longer hospital stays independently form age and sex. The lack of sRD is associated with a greater likelihood to return home from any other state.

The time origin is characterized by a transition into an initial, transient, state, such as the start of treatment; the endpoint is an ‘absorbing’ final transition. Instead of survival data or time-to-event data, data on the history of events is available. Multi-state models provide a framework that allows for the analysis of such event history data. They are an extension of competing risk models, since they extend the analysis to what happens after the first event There are different approaches in the scientific literature trying to assess, at least in part, the landscape of multiple composite outcomes over time. For instance, dynamic prediction of time to a clinical event, that is, the computation of the predictive distribution at a certain moment in time given the history of event(s) and covariates until that moment, as well as of competitive events does not allow studying what occurs after each component of the composite outcome^20,21^.

Multistate Markov Models have "no memory" as these do not consider time durations in the previous states in the calculation of the probabilities of transition to other states.

Currently, new multistate Markov models have incorporated sojourn time in each intermediate state to solve this limitation when this is relevant, that is, the time spent by each individual in each state is now considered. Although these models without memory loss, named semi-Markov multistate models, may be a further advance in this field^22^

There are, however, limitations to this approach. In general, multistate models need big computational efforts for the estimation of parameters, and their estimations are influenced when there are transitions with few observations. The "lack of memory" of multistate Markov Models and the difficulty to solve this limitation has been mentioned before.

## Conclusion

This statistical approach, based on Markov chain models, provides with a statistical solution for the multivariate estimation of the risk of all potential transitions included within mutually exclusive composite endpoints and is a useful complement for complex models of composite outcome analysis, such as the COHERENT model.

## Data Availability

Original data come from the hospital information system. This is not available. Data from the anonimzed edited database may be accessible after reasonable request.

## Non-standard Abbreviations and Acronyms

HF: Heart failure
COHERENT: Clinical Outcomes, HEalthcare REsource utilizatioN, and relaTed costs
CI: Confidence Interval
ED: Emergency department sRD: Severe renal disease

## Acknowledgements

Support statement: This project is an investigator initiative funded by AstraZeneca Farmacéutica Spain, S.A.

## Supplemental Material

Supplementary Table 1 – Supplementary Table 2

